# Triaging of Respiratory Protective Equipment on the assumed risk of SARS-CoV-2 aerosol exposure in patient-facing healthcare workers delivering secondary care: a rapid review

**DOI:** 10.1101/2020.05.13.20101139

**Authors:** Prashanth Ramaraj, Jonathan Super, Ruben Doyle, Christopher Aylwin, Shehan Hettiaratchy

**Affiliations:** Major Trauma Centre, St Mary’s Hospital, Paddington, London, W2 1NY, UK; Department of Engineering, Imperial College London, London, SW7 2BU, UK

**Keywords:** “PPE”, “FFP3”, “surgical mask”, “SARS-CoV-2”, “COVID-19”

## Abstract

**BACKGROUND:** *Objectives:* *“In patient-facing healthcare workers delivering secondary care, what is the evidence behind UK Government PPE Guidance on surgical masks versus respirators for SARS-CoV-2 protection?”*

**METHODS:** Two independent reviewers searched MEDLINE, Google Scholar and grey literature 11^th^ – 30^th^ April 2020. Studies published on any date containing primary data comparing surgical facemasks and respirators specific to SARS-CoV-2, and studies underpinning government PPE guidance, were included. Appraisal was performed using CASP checklists. Results were synthesised by comparison of findings and appraisals.

**RESULTS:** In all three laboratory studies of 14 different respirators and 12 surgical facemasks, respirators were significantly more effective than facemasks in protection factors, reduction factors, filter penetrations, and total inspiratory leakages at differing particle sizes, mean inspiratory flows, and breathing rates. Tests included live viruses and inert particles on dummies and humans.

In six clinical studies, 6,502 participants, there was no consistent definition of “exposure” to determine the efficacy of RPE. It is difficult to define “safe”. The only statistically significant result found continuous use of respirators more effective in clinical respiratory illness compared to targeted use or surgical facemask.

**CONCLUSIONS:** There is a paucity of evidence on the comparison of FRSMs and respirators specific to SARS-CoV-2, and poor-quality evidence in other contexts. Indirectness results in extrapolation of non-SARS-CoV-2 specific data to guide UK Government PPE guidance. The appropriateness of this is unknown given the uncertainty over the transmission of SARS-CoV-2.

1. The evidence base for UK Government PPE guidelines is not based on SARS-CoV-2 and requires generalisation from low-quality evidence of other pathogens/particles.
2. There is a paucity of high-quality evidence regarding the efficacy of RPE specific to SARS-CoV-2.
3. HMG’s PPE guidelines are underpinned by the assumption of droplet transmission of SARS-CoV-2.

Triaging the use of FFP3 respirators might increase the risk of COVID-19 faced by some.

**FUNDING:** This review was unfunded and unsponsored.

**ARTICLE SUMMARY:** *Strengths and limitations of this study:* Strengths

- This article does not aim to prove an intervention as more effective than a comparator. It identifies a paucity of evidence on respiratory protective equipment specific to SARS-CoV-2.
- The results of this study will allow for future study with a real and tangible effect towards the wellbeing of healthcare workers nationwide, and perhaps internationally.
- This article has an exceptionally broad range- from infection control, to public health, to biomechanical engineering, to industry. Its extensive reach would allow for citations from several disciplines. Limitations

- This study reviews evidence specific to a novel virus. Naturally, there is a paucity of specific evidence.

## INTRODUCTION

807 healthcare workers have died of COVID-19 worldwide as of 30^th^ April 2020.^1^ 106 of these tragedies have occurred in the UK.^2^ On 11^th^ April 2020, the World Health Organization (WHO) COVID-19 SitRep^3^ was focused solely on the need for robust reporting of SARS-CoV-2 (the virus causing COVID-19 disease) in healthcare workers (HCWs) in order to better guide infection prevention and control measures.

To have confidence in the indications for use of Respiratory Protective Equipment (RPE),the fluid repellent surgical mask (FRSM) and the Filtering Face Piece Class 3 (FFP3) respirator, UK HCWs must have confidence in the evidence-base behind UK Government (HMG) PPE guidance^i^.^4,5^

It is widely accepted that filtering face piece respirators (that meet UK/EU standards of FFP2/3 and US standards of N95/100^ii^) are more effective in the protection of the wearer from aerosolised pathogens than FRSMs, which are not designed to protect the wearer.^6^ This is reflected in global RPE guidelines ^7–11^ which demonstrate the triaging of respirators to those more likely to encounter aerosolised SARS-CoV-2, and the recommendation of FRSMs to those deemed less likely.

The need for triaging of RPE includes several considerations other than the protective ability of these respirators. These include the shortage of global stock and supply,^7–11^ the need to ensure that low-to-middle income countries (LMICs) are also able to access RPE,^12^ and the relative risk of SARS-CoV-2 exposure by the current understanding of the virus’ transmission.

The latter consideration causes concern. HMG PPE guidance ^4^ on the indications for use of FFP3 respirator relies on two assumptions. Firstly, its list of Aerosol Generating Procedures (AGPs)^5^ and high-risk areas are exhaustive. Secondly, the droplet theory of SARS-CoV-2 transmission ^13,14^ is correct. If either of these two postulates are incorrect and the role of aerosolisation transmission in SARS-CoV-2 is greater than currently thought, the current triaging system of respirators may result in HMG PPE guidance indicating a less effective form of RPE in a higher-than-expected risk setting.

This rapid review aims to determine the evidence-base to the protective ability of respirators versus FRSMs to aerosolised SARS-CoV-2.

## METHODS

This is a rapid systematic review of heterogenous studies with no summary estimate due to vastly different study protocols.

### Review Question^ii^

Following the widely used PICO structure^15^, the research question was framed as:

> “In patient-facing healthcare workers delivering secondary care, what is the evidence behind UK Government PPE Guidance on surgical masks versus respirators for SARS-CoV-2 protection?”

### Preliminary Search for Similar Reviews

Two similar systematic reviews were found.^16,17^ The focus of the Greenhalgh, et al. review ^16^ was on the efficacy of FRSMs and respirators in primary care; while Smith, et al.^17^ did not focus on SARS-CoV-2 prevention, rather respiratory disease in general.

### Search Strategy^ii^

“respirator”, “surgical mask”, “mask”, “FFP”, “FFP3”, “PPE”, “personal protective equipment” AND “viral”, “infection”, “respiratory”, “covid”, “covid–19”, “coronavirus”, “SARS-CoV-2”

Authors PR and JS conducted the following search and eligibility check independently.

### Databases Searched

1. PubMed/MEDLINE.
2. Google Scholar.
3. Grey literature search- by searching for references behind the RPE guidelines of the UK, USA, and EU/EEA^iii^.
4. Snowball search- by reviewing the references of included and excluded articles, and the references of these references, for eligibility and appraisal.

### Eligibility criteria

**Table 1.**
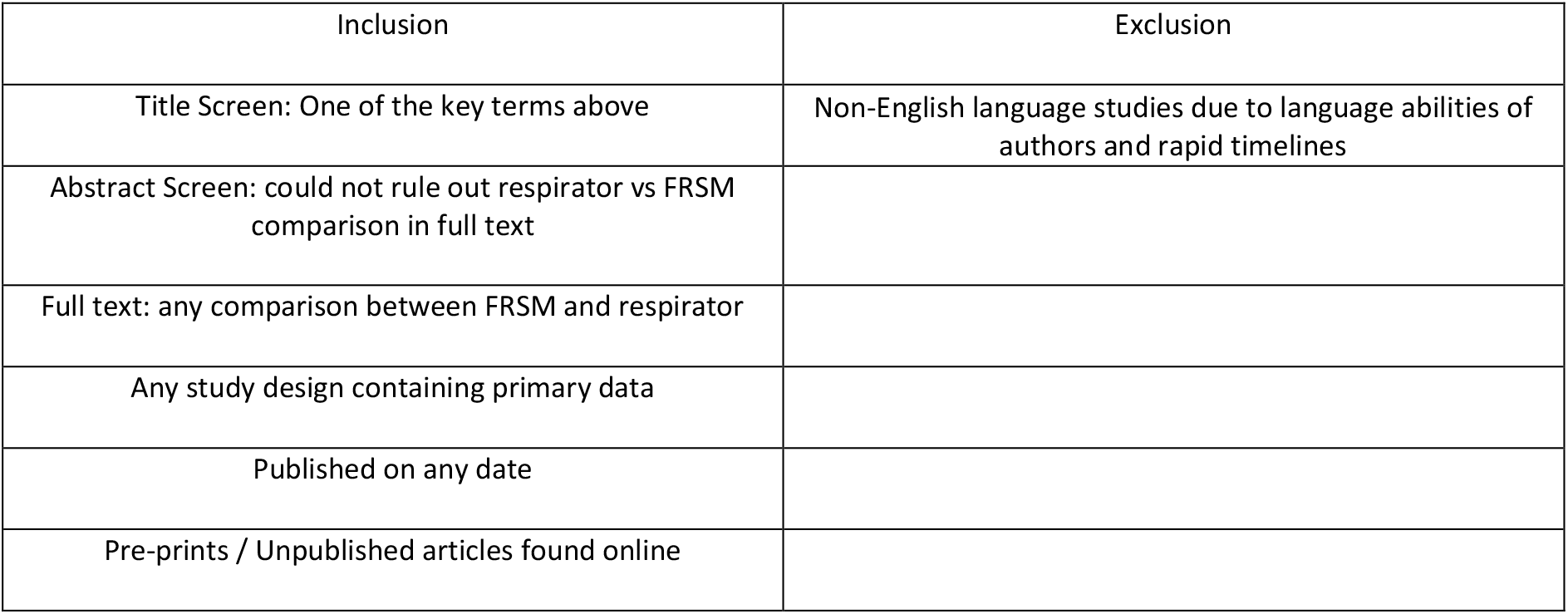
Eligibility criteria for articles discovered through database searching.

### Critical Appraisal

Authors independently used the relevant Critical Appraisal Skills Programme (CASP) checklists.^18^ All studies were included for qualitative analysis since it is noted that during a time of global crisis, the need for rapid evidence based on a novel virus may reduce the viability of gold-standard randomised controlled trials and shorten timelines for follow-up. The need to appraise studies thoroughly for “bad science” is vital during such a time, and therefore comments arising from critical appraisal of all articles included are attached to their results to allow for informed decision making.

### Consensus Meeting

Disagreement resulted in full text review for eligibility and, if accepted, individual appraisals conducted independently. A third author was tasked to review for eligibility had there been any further disagreements on full text review.

### Data Management

A PRISMA ^19^ Flow Diagram can be found on the following page.

### Data Extraction

Data from the 9 included articles were extracted independently onto independent electronic spreadsheets.

Databases were re-searched in the timeframe 11^th^ – 30^th^ April 2020 to identify new literature. An additional similar systematic review was discovered.^20^ This did not contain primary data so was not included, but is discussed as a similar study.

### Result Synthesis

Due to the heterogeneity of study designs and the parameters of results, data extracted from accepted articles were compared directly.

For laboratory studies, these data included study design, research question, masks/respirators tests, testing particle/pathogen, findings, and appraisal comments.

For clinical studies, these data included setting, participants, interventions, outcomes, and limitations raised in appraisal.

### Patient and Public Involvement

This study did not include patient or public involvement due to the rapid nature of the review.

### Data Management

**Figure 1.**
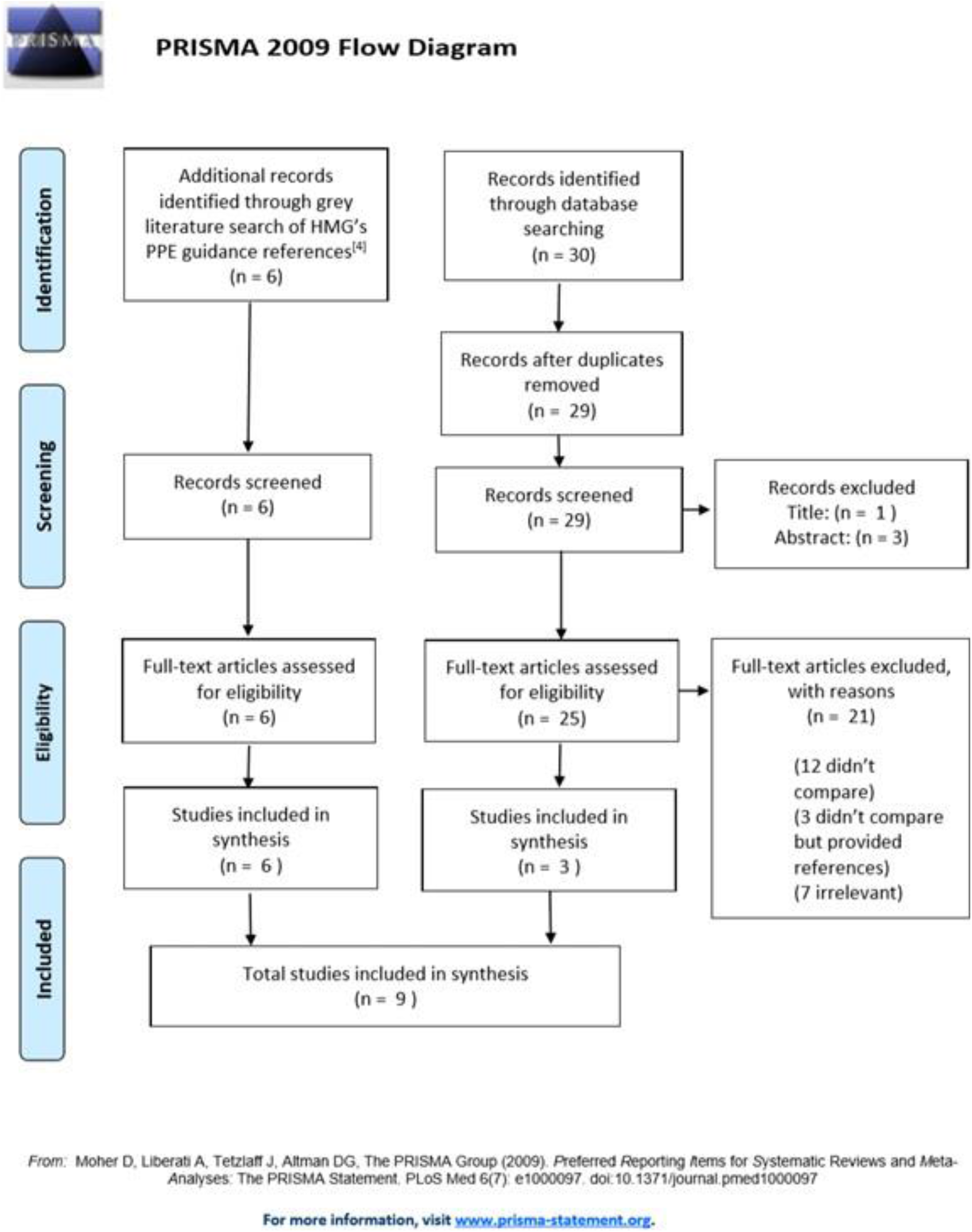
PRISMA Flow Diagram19 displaying data management of search results.

## RESULTS

### Review of laboratory studies comparing respirators with FRSMs

**Table 2.** Results of laboratory studies upon review, illuminating the study design, research question, masks compared, pathogens/particles tested, key findings, and appraisal

| Author | Study Design | Question | Mask/Respirator | Pathogen/Particle | Findings | Appraisal |
| --- | --- | --- | --- | --- | --- | --- |
| Lee SA., et al. 2016.<sup>21</sup> | Human (N = 30) | Do respirators with higher filtration efficiencies provide greater protection when human subjects don the respirators? | 1x FFP2, 1x FFP3 with 3x FRSM | NaCl | 1. The respirators provided between 11.5 to 15.9 times the protection of the FRSMs, suggesting that FRSMs are not a good substitute for respirators when concerns exist about airborne transmission of bacterial and viral pathogens.<br>2. 18.3% of the tested FFP2 respirators had PFs <10, and 41.7% of the tested FFP3 respirators had PFs<sup>1</sup> <<sup>20</sup>, indicating that the European standard for APF of 10 for FFP2 respirators and 20 for FFP3 respirators may overestimate the actual protection offered by these respirators against particles in the size range of 0.093–1.61 µm.<br>3. The protection factors of respirators against particles in the size range of 0.093–1.61 µm were not size dependent. The size ranges of viral and bacterial particles fall into this size range, and they are expected to have similar PFs.<br>4. Correct fit is an important consideration. | + Standardised and peer reviewed method of testing.<br>+ Controlled for age, sex, facial anatomy and hair, fit testing, smoking, previous respiratory use, allergies, cardiovascular/respiratory illness, and drinking within 30 mins of testing.<br>+ Standardised for loss of particles into the sampling devices' lines prior to detection.<br>- No mention of randomisation or blinding.<br>- Small study population (N = 30), narrow age range (18 – 24 year olds), all of Taiwanese origin, respirators from only two companies. Difficult to generalise to wider global population and respirators produced by other companies. |

<sup>1</sup> Protection Factor: a ratio of the test particle/pathogen per unit volume on the outside of the test mask/respirator compared to that on the inside, over a standardised time frame with standardised temperature, humidity, and windspeed.<sup>21</sup>

|  |  |  |  |  |  |  |
| --- | --- | --- | --- | --- | --- | --- |
| Health Safety Laboratory. 2008.<sup>6</sup> | Dummy & Human | What is the contribution of surgical masks in the protection against any residual aerosol risk of airborne particles generated from a simulated sneeze (including those that contain live, infectious influenza virus)? | 11x FFP<br>(2x FFP1)<br>(4x FFP2)<br>(5x FFP3)<br>8x FRSM<br>(5x Tie)<br>(3x Strap) | NaCl &<br>Live *Influenza A* | 1. There is a lack of scientific evidence regarding the protective effect of surgical masks against infectious aerosols (with reference to worker safety) to support HSE's pandemic planning activities.<br>2. Surgical masks will achieve a mean reduction factor<sup>2</sup> of 2 against a simulated sneeze of inert airborne particles.<br>3. The efficiency of FRSMs against inert airborne particles is greatly reduced compared to respirators.<br>4. Live, infectious virus was extracted in enumerable quantities from the air from behind all the surgical masks tested. This suggests that influenza virus can survive in aerosol particles and bypass/penetrate a surgical mask and that a residual infectious aerosol hazard may exist.<br>5. Surgical masks provide a 6-fold reduction in exposure to live, infectious *Influenza A* virus. By contrast, properly fitted respirators provide at least a 100-fold reduction. | + Controlled for *Influenza A* in bioaerosol challenge by calculating reduction factor.<br>+ Standardisation of inert aerosol generation with particle size of human cough.<br>+ Standardisation of fit factors for FFP respirators<br>- No mention of blinding.<br>- Unable to fit FFP respirators to the Sheffield dummy, therefore FFP respirators not tested with live viable *Influenza A*.<br>- Inert testing on one human does not control for different face anatomy<br>- Bioaerosol challenge from only one distance (70cm).<br>- Does not account for environmental factors on *Influenza A* transmission such as humidity, temperature, ventilation. |

<sup>2</sup> Reduction Factor = $\frac{\text{Particle concentration outside the facepiece}}{\text{Particle concentration inside the facepiece}}$

|  |  |  |  |  |  |  |
| --- | --- | --- | --- | --- | --- | --- |
| He X., et al. 2014.<sup>22</sup> | Dummy | How does breathing frequency affect N95 and FRSM performance against viral and other submicron particles? | 1x N95, 1x FRSM | NaCl | 1. The N95 filtered 13.4 times more particles than the FRSM at the highest Mean Inspiratory Flow (MIF) and 108.2 times more particles at the lowest MIF. (N95 $P_{\text{filter}} = 0.72\%$ at 85 L/min (MIF); $0.05\%$ at 15 L/min ($p < 0.0001$). FRSM: $P_{\text{filter}} = 9.65\%$; MIF = 85 L/min; $P_{\text{filter}} = 5.41\%$, MIF = 15 L/min, ($p < 0.0001$)).<br>2. The FRSM allowed the Total Inspiratory Leakage (TIL) of 18.9 times more particles at 10 breaths/minute and 14.9 times more at 30 breaths/min than the N95.<br>N95: (10 breaths/min, mean TIL = $1.22\%$; 30 breaths/min, mean TIL = $1.73\%$ ($p > 0.0025$)).<br>FRSM: (10 breaths/min, mean TIL = $23.1\%$; 30 breaths/min, mean TIL = $25.7\%$) ($p < 0.0025$)). | + measurements controlled for NaCl concentrations higher than environmental concentrations of virus.<br>+ utilised reliable and reproducible parameters.<br>+ controlled for temperature and humidity.<br>+ randomised independent variables to reduce risk of bias.<br>- Only one model of respirator and one mask.<br>- RPE removed after 20 tests, so later tests will be impacted by NaCl loading/"clogging".<br>- RPE taped to mannequin for $P_{\text{filter}}$ testing.<br>- Unclear exactly how many tests were performed. |

### Review of clinical trials comparing respirator with FRSMs

**Table 3.** Results of clinical studies upon review, illuminating the study setting, participant size, masks compared, findings, and appraisal

| Author | Setting | Participants | Interventions | Findings | Appraisal |
| --- | --- | --- | --- | --- | --- |
| Radonovich, et al. 2019.<sup>23</sup> | 7 US medical centres; outpatient setting | 2862 randomized participants | N95 vs FRSM | 1. N95 respirators vs FRSM as worn by participants in this trial resulted in no significant difference in the incidence of laboratory-confirmed influenza | Randomised Control Trial.<br>1. Adherence to infection control was evaluated throughout the study.<br>2. Exposure to patients, co-workers and others with respiratory illness was self-reported in a diary.<br>3. Participants were recruited from the outpatient setting<br>4. Testing methodology - only tested using RT-PCR when symptomatic thus may have missed asymptomatic individuals, as well as 2 random swabs during the study.<br>5. Only assumed that 65% participants were vaccinated against the influenza virus (they did not collect this data) yet they were measuring infection by influenza.<br>6. No protective equipment was worn outside the workplace.<br>7. Only N95 and FRSM masks were tested so authors warn against making generalisations about effectiveness. |
| Loeb, et al. 2009.<sup>24</sup> | 8 hospitals in Ontario, Canada: EDs, AMUs and Paediatric units | 446 nurses | N95 vs FRSM | There were no significant differences between the FRSM and N95 respirator groups in respiratory syncytial virus type B, metapneumovirus, parainfluenza 3, rhinovirus-enterovirus, or coronaviruses.<br>Only 12% of lab confirmed viral infections had fever.<br>No difference between FRSM and targeted N95 use. | 1. The study was abandoned at the start of the 2009 influenza pandemic when all nurses were advised to wear N95.<br>2. Self-reporting of data during 2x weekly questionnaires.<br>3. Only tested if self-reported as symptomatic and sent a test to perform themselves.<br>4. No mention of vaccination history.<br>5. Audits conducted were done via telephone to assess whether patients were admitted to the wards with febrile respiratory illness/influenza. If so, an auditor went into the hospital to observe the use of the protective equipment.<br>6. Only one room entry was recorded per observation.<br>7. Emergency departments were not audited.<br>8. Co-workers and families were not surveyed as a source of infection.<br>9. Hand hygiene, use of gloves and gowns was not monitored. |
| MacIntyre, et al. 2013.<sup>25</sup> | 19 hospitals in Beijing, China: EDs | 1,669 hospital-based workers: nurses, doctors, or ward clerks | Targeted N95 use Vs Continuous N95 use vs FRMS | Rate of CRI (2 or more respiratory symptoms or one respiratory symptom and a systemic symptom) was highest in medical mask (17%) vs targeted N95 (11.8%) and lowest in the continuous N95 arm (7.2%) ($P < 0.05$).<br>Rates of laboratory-confirmed respiratory virus infections were low and not significant between the groups. | Randomised Control Trial.<br>1. Asymptomatic patients were not tested.<br>2. Vaccination status was assessed.<br>3. The study was only carried out for 4 weeks, followed by one week of non-mask wearing to allow for incubation periods.<br>4. Self-reported data using pocket diary (previously validated method of reporting).<br>5. Only conducted for 4 weeks - limitation due to seasonality of different respiratory pathogens. |
| MacIntyre, et al. 2014.<sup>26</sup> | Healthcare workers based in hospitals in Beijing, China | 1441 nurses or doctors, working full time in the emergency department or respiratory wards. | N95 vs FRSM | N95 respirators were significantly protective ($p < 0.05$) against bacterial colonization, co-colonization and viral-bacterial co-infection, compared with FRSM users and the control group.<br>Dual respiratory virus or bacterial-viral co-infections can be reduced by the use of N95 respirators.<br>FRSMs had no significant efficacy against any outcome compared to control. | Randomised Control Trial.<br>1. Participants only tested if symptomatic.<br>2. Participants self-reporting symptoms, hours worked, and masks worn.<br>3. The study was only conducted for 4 weeks.<br>4. No information regarding vaccination history is mentioned.<br>5. Information about potential infection outside of working from co-workers or family was not considered. |
| Ng, et al. 2020.<sup>27</sup> | Inpatient HCWs | 41 HCWs exposed to a COVID-19 positive patient | FRSM vs N95 | None of the health care workers tested positive (PCR) for COVID-19 or experienced any symptoms.<br>85% of the staff were exposed to AGPs whilst wearing FRSM. The other 15% wore N95.<br>There is no evidence that N95 is superior to FRSM. | Case report.<br>1. All patients were swabbed on the same day, ranging from 1-5 days after last exposure to the patient.<br>2. Methodology is poorly reported. Retrospective study design. Small study - only 41 participants with exposure to one single COVID-19 patient. No power calculation.<br>3. Definition of 'exposure' was an AGP of at least 10 minutes, within 2 metres of the patient.<br>6. Conclusion is not based on the results- no evidence to suggest N95 superior to surgical mask does not infer that a N95 and FRSM have the same efficacy. |
| Loeb, et al. 2004.<sup>28</sup> | 2 hospitals in Ontario: coronary care units and ICUs with SARS patients | 43 nurses | Surgical masks vs N95 | Either droplet or limited aerosol generation are the means of transmission to healthcare workers (SARS).<br>Almost 80% reduction in risk for infection for nurses who consistently wore masks (either surgical or N95).<br>When we compared use of N95 to use of surgical masks, the relative SARS risk associated with the N95 mask was half that for the surgical mask; however, because of the small sample size, the result was not statistically significant.<br>Our data suggest that the N95 mask offers more protection than a surgical mask. | Retrospective cohort.<br>1. Use of PPE determined by reviewing documentation and retrospective interviews, may be inaccurate/recall bias.<br>2. No power calculation.<br>3. Subjective measurements of exposure.<br>4. No comment on blinding outcome assessors to exposure (ie did the interviewers know whether the nurse had been SARS +ve?).<br>5. Confounders- any other possible exposure of nurses? Bank shifts at other hospitals? Contact in the break room/canteen?<br>6. Precision of some results questionable- large CIs and p-values (eg in manual ventilation RR). |

## DISCUSSION

### • Statement of principal findings

In all three laboratory studies of 14 different respirators and 12 surgical facemasks, respirators were significantly more effective than facemasks in protection factors, reduction factors, filter penetrations, and total inspiratory leakages for differing particle sizes, mean inspiratory flows, and breathing rates. Both humans and dummies were tested using live viruses and inert particles.

In six clinical studies totalling 6,502 participants, there was no homogenous definition of “exposure” used to determine the efficacy of RPE. Therefore, it is difficult to define “safe”. The only statistically significant result found continuous use of respirators more effective in reducing clinical respiratory illness than targeted use or using surgical facemask.

### • Strengths and weaknesses of the study

A CASP checklist critical appraisal was performed on this review. It was noted that there is a real paucity of evidence regarding RPE specific to SARS-CoV-2. HMG’s PPE guidance ^4^ was found to reference non-SARS-CoV-2 and non-FFP3 specific studies, therefore these were included for review. Any study design containing primary data was included. Non-English language studies were not included, though translated studies were screened. Due to the heterogeneity of study designs, and statistically insignificant results, it was not possible to perform a quantitative analysis. The potential harms of respirator use, such as pressure sores, is poorly documented and requires further study for mitigation and improvement.

### • Strengths and weaknesses in relation to other studies, discussing important differences in results

This review found that just one study directly compares FRSMs and respirators. Ng, et al.^27^ conclude that FRSMs and N95s are equally effective. Limitations of this study include retrospective design, small sample size and a wide range of scenarios defined as ‘exposure’. There is no stratification of confounding variables such as age, sex, health, community exposure to SARS-CoV-2, or exposure to other COVID-19 patients. In the participants tested, none tested positive for SARS-CoV-2 in either intervention. It is unclear how quantitative analysis was performed. No retrospective significant difference was found in SARS-CoV-2 test results of these HCWs. While this does not prove either more effective, it also does not support the study conclusion that FRSMs and N95s are equally effective.

Upon frequent re-searching of literature, a similar systematic review^20^ was published during the study period of this review. This was a systematic review and meta-analysis of four clinical RCTs comparing FRSM and N95 use. Three ^23^,^24^,^26^ of the four studies included in Bartoszko et al.’s review ^20^ were included in this review. That review used search terms specific to RCTs, not specific to SARS-CoV-2/COVID-19, and excluded laboratory studies or tests on mannikins. These authors also highlight the paucity and low-quality of evidence comparing FRSMs and respirators. Their review adjusted for the collation of results from cluster RCTs with individual RCTs. However, the review was not specific to SARS-CoV-2, nor was the meta-analysis of aggregate data specific to any coronavirus. Similarly to this review, that team draw conclusions from laboratory-confirmed illnesses of other viruses to postulate conclusions. Their review might be limited by the exclusion of other study designs, and it is unclear why three studies were included for analysis externally to their search strategy at a late stage, nor why an RCT ^26^ included in this review, providing statistically significant findings, was excluded by that review.

### • Meaning of the study: possible explanations and implications for clinicians and policymakers

There is no high-quality evidence regarding the efficacy of RPE in protecting HCWs against SARS-CoV-2 transmission. There is uncertainty on the transmission mechanism of SARS-CoV-2.^29^ There are challenges to the droplet model of respiratory illness transmission.^30^ Procedures classified as AGPs vary in international guidance.^7–11^ RPE guidance is increasingly stock driven.^8–11^ Given this uncertainty, HMG PPE guidance should take a cautious approach rather than risk under-protecting staff. The evidence base suggests FFP3s may be a more effective form of RPE than FRSM. Due to the current uncertainty surrounding the transmission of SARS-CoV-2, a cautious approach to RPE may be favourable. If RPE must be triaged due to unavailability of stock, FRSM wearing HCWs may be exposed to aerosolised SARS-CoV-2.

### • Unanswered questions and future research

Further rigorous study is required into the transmission of SARS-CoV-2, as recent studies liken it more to SARS-CoV–1 than to influenza. HMG PPE guidance is based off preparedness for an influenza pandemic.

The validity of the droplet vs aerosol dichotomy of respiratory illness transmission is uncertain. It must be substantiated since it underpins HMG PPE guidance on RPE.

Expedited research is required to further understand aerosol-generating procedures, including an effort to homogenate the classification of AGPs by different organisations as AGPs are the key indication for RPE triaging in HMG PPE guidance.

HMG PPE guidance on the indications for use of FFP3 and FRSM is underpinned by the droplet theory of transmission of SARS-CoV-2, based on the flowchart below suggested by Coia, et al. in 2013.^31^

**Figure 2.**
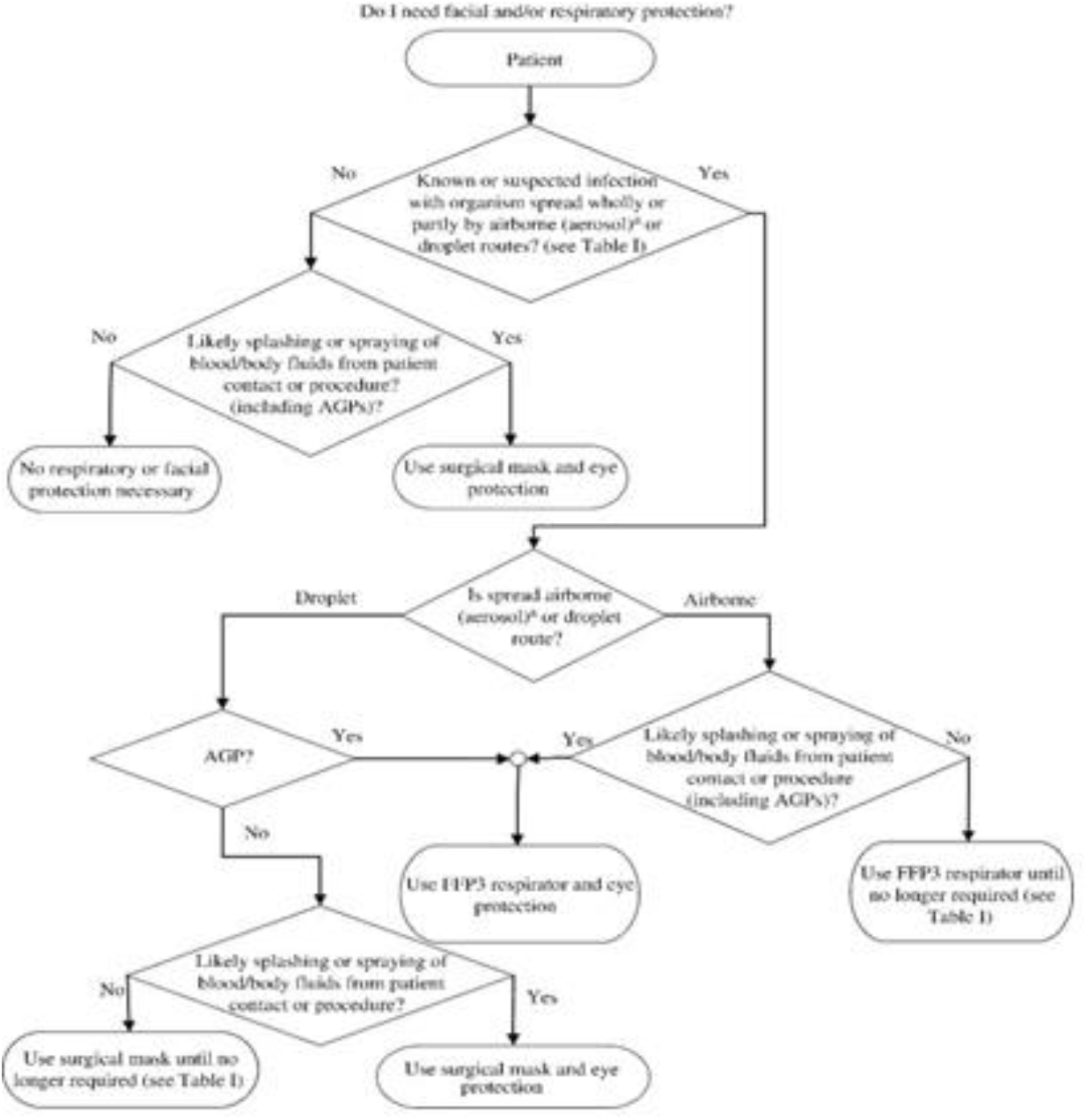
Flowchart designed by Coia et al31 underpinning HMG PEE guidance.^4^

## CONCLUSION

HCWs away from work, self-isolating, or on sick leave due to COVID-19 reduce the health system’s capacity to deal with the ongoing pandemic ^3^. In order to reduce sickness burden on health systems, HCWs must be able to make informed, evidence-based decisions on their choice of PPE.

This review concludes:

1. The evidence base for HMG’s PPE guidelines ^4^ is not based on SARS-CoV-2 and requires generalisation from low-quality evidence in which other pathogens/particles were tested.
2. There is a paucity of high-quality evidence regarding the efficacy of RPE specific to SARS-CoV-2.
3. HMG’s PPE guidelines are underpinned by the assumption of droplet transmission of SARS-CoV-2.

It is evident from WHO ^32^, ECDC^9–11^, and CDC^8^ guidance that the indications for the use of RPE are not based solely on the protective abilities of respirators and FRSMs. Instead, a triaging system based on an expected shortage of global stock and supply, combined with current understanding of likelihood of exposure to aerosolised SARS-CoV-2 is used.

There is active discussion regarding the droplet transmission of SARS-CoV-2 with an accepted uncertainty in understanding. Given this uncertainty, a cautious approach should be taken in the protection of HCWs. This review found that in all laboratory studies respirators were more protective to the wearer than FRSMs in all parameters tested. In the clinical studies reviewed, the only statistically significant finding was that respirators provided significant protection against bacterial-viral coinfection compared with FRSMs. No statistically significant evidence was found to support the conjecture that an FRSM might provide the same level of protection as a respirator against SARS-CoV-2, or indeed any tested live virus or inert submicron particle. Therefore, use of a respirator would be the more cautious option.

While the triaging of RPE due to a lack of global stock is understandable and appropriate during the strains of a pandemic, it must be noted that by increasing the protection of some through the provision of respirators, HMG PPE guidance might be increasing the risk of COVID-19 faced by others.

## Data Availability

The authors will support data sharing on request by emailing the corresponding author, PR.

## AUTHOR STATEMENT

### Author Contributions

All authors meet ICMJE’s four criteria of authorship. PR conceived the idea, contributed to the methodology, conducted the search, extracted and appraised data, and wrote the article. JS contributed to the methodology, conducted the search, extracted and appraised data, and reviewed the article. RD contributed to the writing of the article, and provided review and guidance throughout. CA contributed to the writing of the article and provided review and guidance throughout. SH contributed to the development of concept, contributed to the writing of the article and provided review and guidancethroughout. All authors approved the final manuscript and article submission.

#### Acknowledgements

The authors would like to acknowledge the following for their peer review of the completed manuscript prior to submission:

Mr Isaac Florence MSc

Dr Rory Wilson MBBS BSc (Hons), University of Glasgow School of Medicine, Glasgow, G12 8QQ Miss Roseanna Jenks RM MA (Cantab), West Middlesex University Hospital, London, TW7 6AF

#### Guarantorship

The corresponding author attests that all listed authors meet authorship criteria and that no others meeting the criteria have been omitted.

#### Copyright

The Corresponding Author has the right to grant on behalf of all authors and does grant on behalf of all authors, a worldwide licence to the Publishers and its licensees in perpetuity, in all forms, formats and media (whether known now or created in the future), to i) publish, reproduce, distribute, display and store the Contribution, ii) translate the Contribution into other languages, create adaptations, reprints, include within collections and create summaries, extracts and/or, abstracts of the Contribution, iii) create any other derivative work(s) based on the Contribution, iv) to exploit all subsidiary rights in the Contribution, v) the inclusion of electronic links from the Contribution to third party material where-ever it may be located; and, vi) licence any third party to do any or all of the above.

#### Transparency and Ethics Approval

The lead author (the manuscript’s guarantor) affirms that the manuscript is an honest, accurate, and transparent account of the study being reported; that no important aspects of the study have been omitted; and that any discrepancies from the study as originally planned (and, if relevant, registered) have been explained. Ethics approval was not required.

#### Data Statement

The authors will support data sharing on request by emailing the corresponding author, PR.

## Footnotes

i Please see Appendix 1 for HMG PPE guidance.

ii Please see Appendix 2 for a comparison of the various international standards of testing of respirators and surgical facemasks.

iii Bodies outside of the UK were searched since it was felt that these populations have similar demographics and pandemic response measures.

iv Full PICO strategy, and search strands available in Appendix 3.

